# Stakeholder Perspectives on Long-Acting Injectable PrEP and MOUD: A Descriptive Qualitative Study

**DOI:** 10.64898/2026.06.25.26356527

**Authors:** Luzan JadKarim, Hannah Camp, Aurielle Thomas, Thomas Blue, Michael Gordon, Curt G. Beckwith, Melissa J. Zielinski, Jennifer L. Vincenzo, Lauren Brinkley-Rubinstein

## Abstract

Long-acting injectable (LAI) formulations of pre-exposure prophylaxis (PrEP) and medications for opioid use disorder (MOUD) offer promising new approaches for addressing the intersecting challenges of HIV and opioid use disorder (OUD) in carceral settings. However, successful implementation requires understanding the perspectives of key carceral and community stakeholders. This qualitative study involved 17 semi-structured stakeholder interviews including correctional staff (medical and administrative) and community healthcare providers. Participants were recruited from jails in Washington, DC and surrounding Maryland counties. Interviews were coded using a mixed inductive and deductive methodology, and data were analyzed to identify key themes. Four overarching themes relevant to implementation emerged: (1) The presence of resource barriers, including jail staffing shortages and financial constraints; (2) Challenges administering medications including stigma, medical mistrust, and limited patient knowledge of medications; (3) challenging and restrictive carceral infrastructure; and (4) the importance of care continuity during carceral-to-community transition. Stakeholders emphasized that while LAI PrEP and MOUD could provide critical support to justice-involved individuals, systemic barriers challenged successful implementation and delivery in jail and during community re-entry. Our findings underscore the need for targeted interventions to address the resource, medication, and infrastructure challenges identified. Ensuring sustainable funding, providing staff training and education, improving patient education, addressing stigma, and strengthening community linkages are essential for the successful implementation of LAI PrEP and MOUD in carceral settings. These insights can inform future policies and practices aimed at improving health outcomes for justice-involved populations.

## Introduction

People who are incarcerated have much higher rates of HIV and opioid use disorder (OUD) than those in the general population[1–5]. Each year, nearly eleven million people pass through jails in particular—spaces characterized by rapid turnover and limited access to healthcare. Unlike prisons, jails in the US refer to local or state-operated correctional facilities that detain individuals awaiting trial, sentencing, or those with short sentences [6]. Individuals with a history of incarceration are disproportionately impacted by substance use disorders and face heightened risks for HIV, making jails a crucial setting for preventive and therapeutic interventions[2, 7].

The Optimizing Long-Acting Pre-Exposure Prophylaxis and Medications for Opioid Use Disorder Interventions in Carceral Settings (NOTRE) effectiveness trial seeks to prevent and reduce fatal overdose (OD) through the development, implementation, and evaluation of a tailored treatment protocol involving long-acting injectable (LAI) formulations. LAI formulations for pre-exposure prophylaxis (PrEP) and medications for opioid use disorder (MOUD) offer an innovative and potentially relevant medication delivery method for persons in jails. Unlike daily oral medications, which are the primary options in carceral facilities, LAIs can provide sustained protection and treatment alleviating concerns about daily adherence. In a study exploring long-acting antiretroviral therapy (LA-ART) among people living with HIV who use drugs, Chayama et al.[8] found that people at risk of HIV valued the reduced pill burden and greater privacy offered by injectables [9]. Similar findings have emerged for long-acting MOUD. Thepmankorn et al. [7] reported that while many individuals with opioid use disorder were unaware of extended-release buprenorphine (XR-BUP), those who had used it described it as a stabilizing force, protecting them from daily cravings and the chaos of street drug use.

Individuals with SUD face 40 to 129 times the rate of fatal OD within the first two weeks following release when compared to the general population [1, 10, 11]. And while the risks of fatal OD post-release are well documented, the risks of nonfatal OD are more ambiguous despite their common occurrence once incarcerated individuals are back in community [11]. Regardless of the type of OD, individuals released from incarceration face a disproportionate risk of experiencing relapse, OD and death. Therefore, treatment in the form of LAI formulations can be a lifesaving option for individuals nearing release from incarceration.

However, even as LAIs promise to transform care, carceral environments are often shaped by stigma and structural issues that may hinder their implementation [12]. Limited staffing, restricted medical resources, high costs of LAI?, and rapid turnover of incarcerated persons in jails complicate consistent medication administration [12]. Moreover, stigma—both institutional and interpersonal—can discourage individuals from accessing HIV and OUD treatments, while logistical challenges, such as maintaining proper storage conditions for injectables, further hinder implementation.

Therefore, to design and implement a successful protocol for the NOTRE trial, we sought to capture the perspectives of stakeholders directly involved in the future implementation of co-located LAI PrEP and MOUD in large metropolitan jails. By understanding their insights, we aim to design a program that is not only clinically effective but also contextually appropriate to support implementation.

## Methods

### Study Design

NOTRE aims to develop, implement, and evaluate an intervention protocol to deliver LAI PrEP and MOUD to incarcerated individuals around the time of release. We interviewed key stakeholders as part of the first aim of NOTRE to gain carceral and community perspectives on the protocol development.

### Stakeholders

We interviewed 17 stakeholders in Washington, DC, and surrounding Maryland counties representing carceral staff (medical and administration) and personnel from community health organizations [13, 14]. Both carceral staff and personnel from community-based organizations were enrolled to reflect the continuum of care from incarceration through community re-entry. Participants were recruited from a convenience sample of individuals who worked at or with the jails in DC and surrounding Maryland counties and through referrals. Interviews were conducted virtually and participants provided verbal informed consent for study participation and to be recorded electronically.

### Data collection

Semi-structured interviews were conducted by a researcher trained in qualitative interviewing, with each session lasting approximately one hour. We used a semi-structured interview guide to ensure consistency while allowing flexibility for participants to share their perspectives freely. Questions from the interview guide are included in table 1 of the appendix. Interview questions focused on the following general themes: 1) participants’ knowledge and attitudes toward LAI PrEP and MOUD, 2) anticipated barriers and facilitators to LAI implementation in jail settings, and 3) recommendations for optimizing the LAI interventions in jail and the community settings after release. All interviews were audio-recorded and transcribed by a professional transcription service.

**Table 1.**
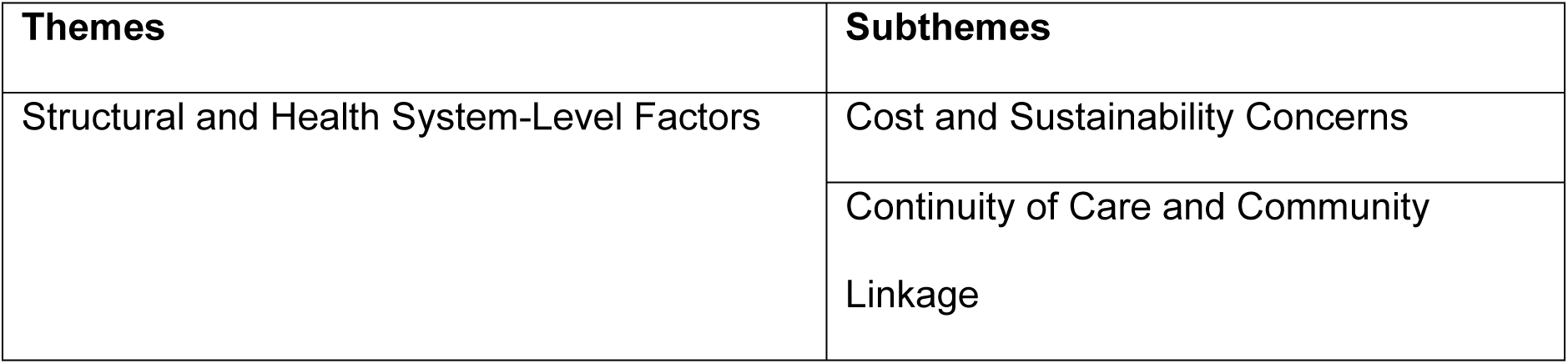
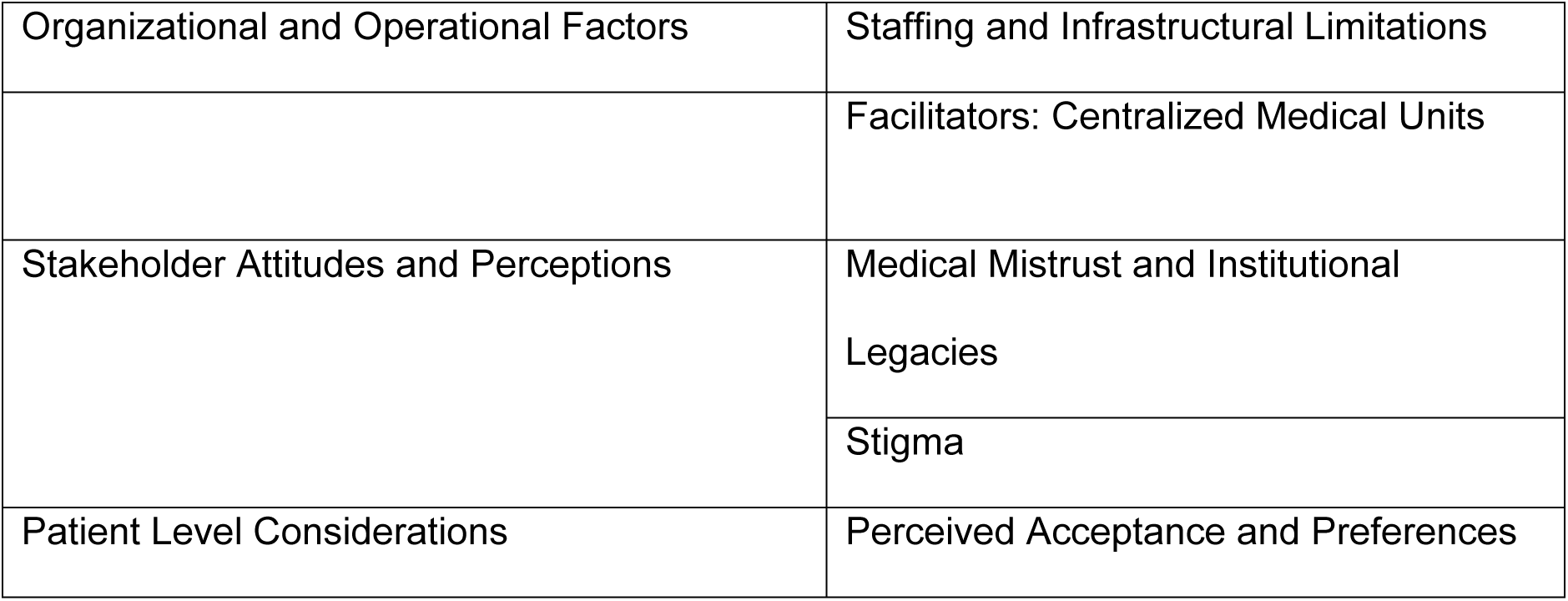
Themes and subthemes.

### Data analysis

We used a mixed inductive and deductive approach to create the codebook from our interviews focusing on key themes. Two researchers with training in qualitative interviewing and analysis independently coded each anonymized interview in full using NVivo14[15] and met to discuss discrepancies with the code. Once the transcripts were coded with high agreement, the two researchers identified themes and subthemes that reflected the stakeholders’ experiences with administering and engaging incarcerated individuals with treatments in pre-and post-release. Quotes that described each subtheme were then selected to support data interpretation and illustrate the barriers and facilitators of the use of LAI formulations both within carceral facilities and in communities.

### Ethical considerations

This study protocol was reviewed and approved by the Duke Institutional Review Board (Pro00115412). The research team conducting this study were not related to any of the participants interviewed and represent academic research institutions that are unrelated to any carceral system and/or community healthcare organization. Verbal and written consent were obtained from all participants in the study.

## Results

### Demographics

Of the 17 participants interviewed, 11 identified as men and six identified as women. Ten participants were White, five participants were Black, and two participants identified with another racial background. Our stakeholders also represented a wide range of professions across carceral settings, and included correctional staff (wardens, directors, and inmate services) and medical staff (therapists, nurses, and substance use coordinators).

### Qualitative Results

There was enthusiasm and acceptance of LAI formulations across our participant population. Every participant expressed their support in implementing the formulations and the promise they offer to both incarcerated and released individuals with OUD and high risk of HIV. However, a common theme among all participants was the need to ensure optimal implementation that addresses barriers and leverages facilitators. Stakeholder interviews revealed a complex set of structural, organizational, and interpersonal dynamics shaping the implementation of LAI PrEP and MOUD in carceral settings. Barriers and facilitators clustered around four key domains: (1) structural and health system-level factors, (2) organizational and operational constraints, (3) stakeholder attitudes/perceptions; and (4) patient-level considerations. While many stakeholders saw potential for innovation and improved outcomes with long-acting formulations, concerns around cost, sustainability, staffing, stigma, and patient trust presented significant implementation challenges.

### Themes

Table 1 presents the four key themes that emerged from participant interviews. Each theme is further broken down into multiple subthemes.

### Structural and Health System Level-Factors

Most participants noted that despite their support for LAI PrEP and MOUD, they had cost and sustainability concerns regarding its provision and distribution. LAI formulations of PrEP and MOUD are expensive, and participants indicate that most jails cannot afford to offer the drug without specific grants as one participant stated,

> *“Yeah, absolutely. The problem being cost prohibition and no Medicaid involvement while somebody’s in the jail. So, our hands are frequently tied. We have a great number, a growing number of people that are on either BRIXADI or SUBLOCADE right now in some part of the jail.”*

Brixadi® and Sublocade® are two LAI formulations for MOUD, and several participants have named their prohibitive expensiveness. Another participant even said, *“… cost is definitely a concern because we actually have a big barrier right now only because we have doses for Sublocade, but it’s only for ten patients because we had a grant with our program that supplied us enough injectables for ten patients.” I think right now we’ve only maybe had three people on the injectable, and the thing is we can’t ask for more money because [county funders] don’t see evidence of it being used.”*

Furthermore, stakeholders perceived that decision makers often only see the immediate costs rather than the long-term savings. Several participants noted that LAI formulations, while more expensive in the short-term, may result in long-term savings, but policy makers often view this using a short-sighted lens. One participant specifically highlighted that LAI medication *“…saves everything,”* and that “… *it saves money on the cost that you’re spending on custody and security watching them* [incarcerated individuals] *do this stuff and all.”*

Relatedly, multiple participants emphasized the growing interest of LAI PrEP and MOUD among incarcerated individuals but had concerns about long-term sustainability of funding. As noted above, often grants could be obtained to cover the costs of medications, but there were concerns about what would happen once those funds were gone:

> *“ … there needs to be continual – doing something for the good to see if it works, it needs to have a longer plan of sustainable funding and support by legislature as well as county council, so this isn’t a population that – I get very guarded when we wanna try something new but it ends, and that’s very frustrating. It’s frustrating because if we mean this is where it should be and this is where it is, that buy in has to be at the highest levels.”*

The transition from jail to community care represents a critical point of vulnerability and a breakdown in continuity of care. Many facilities lack formalized processes for linking individuals to ongoing treatment, and individuals are at a heightened risk of losing their access to medications and healthcare after release. One participant emphasized the importance of continuity of care, explaining that *“for some of our highest-risk clients, having the injection on board when they leave gives us peace of mind. It reduces their risk of overdose and HIV exposure during the chaotic first month post-release.”*

Several participants underscored the critical importance of LAI PrEP and MOUD as a harm reduction tool for individuals transitioning from incarceration to the community. By administering an LAI before release, healthcare staff can be confident that their most vulnerable clients are leaving the jail with a critical layer of protection, even if they struggle to maintain consistent care in the community. This proactive approach helps bridge the gap between carceral and community care, transforming the release process from a moment of extreme vulnerability to an opportunity for sustained protection. However, when the injection is not available, many individuals are at risk of experiencing treatment gaps.

Strong community partnerships can mitigate these risks. As one participant noted that having a provider within close proximity to where released individuals are staying can “…*increases the probability that they will maintain their compliance.”* This strategy emphasizes the importance of familiar, accessible providers who can offer consistent care, especially for individuals who are at the highest risk of loss to follow up immediately following release. Another participant highlighted the impact of re-entry case management,

> *“Our re-entry case managers give people direct contacts and resources before they leave, which increases the likelihood they will maintain care post-release.”*

Such proactive planning transforms the point of release from a moment of risk into an opportunity, ensuring that individuals have a clear path to continue their care. Effective re-entry strategies offer a bridge between the carceral and community health systems, increasing the likelihood of sustained treatment adherence.

### Organizational and Operational Factors

Jail environments are marked by constant turnover and limited continuity of care, creating significant challenges for medical interventions. Participants consistently described how rapid turnover, high population mobility, and limited staffing create substantial barriers to consistent care. One participant noted,

> *“Jail is chaotic, and security rather than healthcare is the priority. More often than not, it takes days for individuals to identify themselves as being HIV infected, have medications confirmed, and receive their ART.”*

While this example was for HIV treatment, such conditions delay many essential treatments. This ultimately highlights how slow processes, coupled with a lack of medical prioritization, leave vulnerable individuals without timely access to care. This instability is further compounded by sudden releases, as another participant explained, *“…a lot of people get released suddenly in court, and we lose them before we can set them up with care.”*

Such sudden releases are not just a logistical problem, but a structural one—where the unpredictability of the justice system directly disrupts healthcare continuity. Without sufficient pre-release planning, individuals may be released without necessary medications or a clear plan for follow-up care.

In addition to infrastructural limitations, multiple members of both carceral and community-based clinics noted the lack of staffing, specifically nurses. One participant stated that, *“…in the end, it’s staffing, it’s staffing, it’s staffing, and no medical mandate or great suggestion or good idea.”* They went on to say that they “*… run an operation that goes on beyond the normal business hours, and so I know what I can do versus I know sometimes, hey, we’d like to do this and that. Hey, I’d love to. How do you wanna do it? There’s a national nurse shortage, and I’m short about five, which is about 22% of my total complement”*

Nurses are the “backbone” of the medical operations within the carceral setting and in order to implement LAI formulations, more nurses need to be onboarded. Multiple participants voiced their concerns with the current nurse shortage as a barrier in addition to the costs required to transition from oral to LAI formulations.

Despite certain limitations mentioned above, infrastructure can also act as a facilitator. Jails with centralized medical units provide a structural advantage. As one staff member highlighted, “Having all MAT patients in one unit helps provide treatment and reduces diversion risks.” Another described how a designated medical unit ensures continuity of care, noting,

> *“Our jail has a designated medical unit with staff that ensures continuity of care. That structure helps us manage treatment better.”*

Such configurations streamline care and enhance patient outcomes. Participants at each of our partner sites have named this as a strength. It makes sense logistically, reduces the amount of time it takes to distribute medications and coordinate medication delivery, and several participants also noted its potential for reducing stigma and enhancing peer to peer support, especially around substance use.

### Stakeholder Attitudes and Perceptions

An overarching barrier to LAI PrEP and MOUD that surfaced was medical mistrust. It is a well understood phenomenon that carceral settings reflect a higher proportion of certain minority groups compared to the general population. Throughout the interviews, numerous participants prefaced their statements regarding medical mistrust in jails with the history of research among these minority populations. As a result, the population has grown wary of newer medical treatments. One participant specifically said that their *“…chief health officer at the time, in a closed door meeting with just the higher up leadership, kind of broke down in tears talking about the mistrust that he learned from his family because of the stuff like the Tuskegee experiment. And that they couldn’t trust stuff coming out of the government, they were being a test subject of some kind.”* The participant then went on to elaborate further on the mistrust in research stating that incarcerated individuals had “*… just a general mistrust of correctional agencies.”* They went on to describe the introduction of a program implemented years ago named SafeLink, also known as “The Obama Phones”, which sought to connect incarcerated individuals leaving jails with “…*a free phone. And we actually had one of the companies providing those phones set up in our lobby on certain days to give out the phones. People wouldn’t take them.”* Some participants also expressed that the carceral setting amplifies this medical mistrust, with one denoting that,

> *“.. this kind of setting, the setting itself, is just a negative setting. And, sometimes people come in with a preconceived notion that I’m in jail. These people are just gonna treat me poorly, and then you have to kind of break that thought process. And then, you do get bad experiences from people, so I’m really big on being responsive and showing that you care to the patient population.”*

In addition to medical mistrust, participants discussed stigma surrounding PrEP and MOUD among their populations. Most participants specifically highlighted the stigma associated with PrEP as an HIV prevention effort. Specifically, one participant stated that, *“the only area I can say that’s a disadvantage is some people may be a little reluctant to wanna go with the HIV prevention only because of the stigmatization of it, and particularly the men population, they don’t like to acknowledge anything that’s wrong. Medicine might make me justify that something’s wrong with me or something, those kinds of stigmatizations.”* He also discussed issues with homophobia detailing that “*…inmates don’t want anyone thinking that they’re vulnerable, that they identify as gay.”* This perspective was echoed by the majority as stigma surrounding HIV is higher among incarcerated individuals, with one participant expressing that,

> *“…I believe that it’s related to the way our population has been treated in medical settings for a very long time. They’ve learned to hide things because they’re judged for them, and it’s about the stigma about having a substance use disorder, the stigma about having multiple partners or having sex with the same sex. I just think they’ve been treated so poorly in the traditional healthcare system that when we get them, we try to just be so nonjudgmental because we are nonjudgmental.”*

Despite the aforementioned stigma, participants at each of our sites continued to voice their positive perspectives surrounding LAI formulations. Several even mentioned that the privacy and confidentiality granted to receiving an injectable may help reduce the stigmatization that surrounds HIV prevention and treatment.

### Patient Level Considerations

While participants held general positive attitudes towards LAI formulations, they expressed multiple downsides. One barrier to the implementation of LAI PrEP and MOUD mentioned was the fear of needles. Some participants noted that people who are incarcerated may fear or avoid injections. One participant specifically mentioned that,*“…sometimes they don’t like to get shots. People just don’t… They’re afraid of needles or whatever.”* Participants also expressed concern that acceptability may be lower without proper education or reassurance. One participant mentioned,

> *“But, then there are the other folks that it’s gonna take them a little while to trust us, and we’re okay with that. We’ve gotta build that trust. We have to earn it…I guess for us it would really be about making sure that the staff had all the education they needed around this so they could adequately prepare the patient and give them all the information that they needed to do this.”*

However, all participants reiterated their support for LAI medications despite these barriers. Many also mentioned the growing interest in the medication among the incarcerated population. One participant compared LAI to daily medications citing its flexibility and advantages,

> *“And so, I think there’s just a ton of advantages to one and done. And I love the flexibility of BRIXADI that you can do these shorter term or shorter duration injections…daily oral can get lost. Daily oral can get stolen. Daily oral can be left on the bus.”*

Perspectives from participants in both the clinical and jail settings were also positive, with one participant indicating that,

> *“…I would support it, and again, based on a screening to see if it’s long-term appropriate for this particular individual… I think it would be very advantageous for them, but always in conjunction with, as you said, the counseling around HIV, relationships, behavior, as well as the treatment aspect, the therapy around the opioid use.”*

Despite the stigma and medical mistrust accompanying the medications, participants indicated their readiness to implement LAI PrEP and MOUD in both carceral and community settings.

## Discussion

Our study explored the perspectives of stakeholders on the implementation of long-acting injectable (LAI) PrEP and MOUD within carceral settings. The findings revealed a complex landscape marked by substantial barriers and facilitators across four major themes: structural and health system-level factors, organizational and operational factors, stakeholder attitudes and perceptions, and patient level considerations. Our results highlight both the promise and challenges of implementing LAI interventions in these high-risk settings.

Participants consistently identified staffing, cost, and continuity of care as critical structural and organizational barriers. The national nursing shortage, coupled with the unique demands of jail settings, exacerbates staffing challenges, making it difficult to ensure that LAI medications are administered effectively[16]. Cost was another significant concern, with stakeholders expressing frustration over the lack of sustainable funding to support LAI PrEP and MOUD. These findings align with existing literature highlighting the financial and human resource constraints in correctional healthcare settings [12]. While not surprising, the lack of resources to introduce a treatment like LAI is a gap where research studies like NOTRE can fill. However, while trials can introduce much needed interventions, researchers should also consider maintaining the intervention’s accessibility and sustainability post-trial. Our study further emphasizes the importance of ensuring sustainable funding and workforce development to make LAI options viable in these environments.

Stigma and medical mistrust emerged as central barriers to patient uptake of LAI PrEP and MOUD. Participants described how stigma surrounding HIV and substance use can prevent individuals from accessing these interventions, a well-established barrier in prior research [17, 18]. Medical mistrust, particularly in communities with histories of exploitation, further complicates the implementation of novel treatments like LAI [7, 18, 19]. These insights are consistent with prior research, which has documented the role of stigma and mistrust in limiting healthcare access for justice-involved populations [7, 19, 20]. These barriers provide valuable insight for the NOTRE trial as one of its main objectives is a sustainable implementation model for LAI medications. Through tailored education, establishing rapport with participants, and demonstrating connections with community providers and resources, NOTRE aims to overcome the stigma and medical mistrust described by our participants. Furthermore, the upcoming trial has and will continue to receive input from the NIH-funded INTERACT (Using DesIgN jusTicE to impRove cArCeral health ouTcomes) Center that will inform best practices from persons with context expertise in HIV, SUD, and CLI for the sustainable implementation of LAI medications in the carceral setting.

The chaotic nature of carceral settings creates significant challenges for consistent care delivery. Our findings revealed that without robust medical infrastructure, it is difficult to maintain continuity of care. These structural barriers are well-documented in the literature [2, 3], yet our study highlights how they directly impede the implementation of LAI interventions.

Participants underscored the critical role of the carceral-to-community transition, with LAI PrEP and MOUD viewed as key tools for helping to maintain care continuity post-release. The medication provides consistent medical level between doses to bridge the gap in care post-release. However, gaps in community coordination and follow-up threaten the success of these interventions. Our findings echo previous studies that emphasize the importance of strong community partnerships and re-entry planning to support individuals transitioning from incarceration [12].

Our findings offer several implications for practice. First, stakeholders must advocate for sustainable funding mechanisms to ensure the long-term availability of LAI PrEP and MOUD, such as advocating for 1115 Medicaid waivers. Also known as Medicaid reentry waivers, these waivers allow Medicaid to cover certain services for individuals prior to their release from incarceration [21]. These state-initiated waivers have the potential to reduce overdose mortality, emergency room visits, and other health outcomes post-release by bridging the gap in care upon release [21]. Second, targeted education for staff and patients can help mitigate stigma and build trust. Third, jails should be supported in developing robust infrastructure to accommodate LAI administration, including dedicated medical units and streamlined procedures, and working collaboratively with pharmacy. Finally, strong partnerships with community providers are essential to support individuals as they transition from jail to the community.

### Limitation

Our study’s findings should be interpreted with some limitations to consider. First, our data was collected from different county locations, meaning the perspectives collected are specific to those jurisdictions only. Second, despite efforts to minimize bias, data collection and interpretation were inevitably shaped by the researchers’ perspectives and should be considered in that context.

## Conclusion

Our study provides important insights into the perspectives of stakeholders regarding the implementation of LAI PrEP and MOUD in carceral settings. Addressing the identified barriers—particularly those related to resources, stigma, infrastructure, and re-entry planning—is critical for the success of these interventions.

## Data Availability

The data underlying this study consist of semi-structured interview transcripts containing sensitive information provided by our participants. Participants were not asked to consent to public data sharing, and full public disclosure of interview transcripts could compromise participant confidentiality. Accordingly, the raw data cannot be made publicly available. De-identified data supporting the findings of this study are available upon reasonable request to the corresponding author.

## Conflict of Interest

The authors of this manuscript do not have any conflicts of interest to disclose.

## Notes

**Funding:** This research was supported by NIH grant R61DA060583

### Competing Interest Statement

The authors have declared no competing interest.

### Clinical Trial

NCT06854029

### Funding Statement

Yes

### Author Declarations

This study protocol was reviewed and approved by the Duke Institutional Review Board (Pro00113660).

## References

1. Binswanger IA, Krueger PM, Steiner JF. Prevalence of chronic medical conditions among jail and prison inmates in the USA compared with the general population. J Epidemiol Community Health. 2009;63(11):912–9.

2. Blue C, Buchbinder M, Brown ME, Bradley-Bull S, Rosen DL. Access to HIV care in jails: Perspectives from people living with HIV in North Carolina. PLoS One. 2022;17(1):e0262882.

3. Levano SR, Epting ME, Pluznik JA, Philips V, Riback LR, Zhang C, et al. HIV testing in jails: Comparing strategies to maximize engagement in HIV treatment and prevention. PLoS One. 2023;18(6):e0286805.

4. Shook-Sa BE, Hudgens MG, Kavee AL, Rosen DL. Estimating the Number of Persons with HIV in Jails via Web Scraping and Record Linkage. J R Stat Soc Ser A Stat Soc. 2022;185(Suppl 2):S270–s87.

5. Maruschak LM. HIV in Prisons, 2021 – Statistical Tables. U.S. Department of Justice: Office of Justice Programs - Bureau of Justice Statistics; 2023.

6. Sawyer W, Wagner P. Mass Incarceration: The Whole Pie 2025. Prison Policy Initiative; 2025.

7. Thepmankorn P, Flumo R, Nyaku AN. Perceptions of People Who Inject Drugs About Long-acting Medications for Opioid Use Disorder, Preexposure Prophylaxis, and Antiretroviral Therapy. Open Forum Infectious Diseases. 2025;12(3).

8. Chayama KL, Ng C, Brohman I, Mansoor M, Small W, Philbin M, et al. Acceptability of long-acting antiretroviral therapy among people living with HIV who use drugs in Vancouver, Canada: A qualitative study. PLOS ONE. 2025;20(2):e0319010.

9. Collins AB, Macon EC, Langdon K, Joseph R, Thomas A, Dogon C, et al. Perceptions of Long-Acting Injectable Antiretroviral Therapy Among People Living with HIV Who Use Drugs and Service Providers: a Qualitative Analysis in Rhode Island. J Urban Health. 2023;100(5):1062–73.

10. Balter DR, Howell BA. Take-Home Naloxone, Release From Jail, and Opioid Overdose—A Piece of the Puzzle. JAMA Network Open. 2024;7(12):e2448667-e.

11. Hartung DM, McCracken CM, Nguyen T, Kempany K, Waddell EN. Fatal and nonfatal opioid overdose risk following release from prison: A retrospective cohort study using linked administrative data. J Subst Use Addict Treat. 2023;147:208971.

12. Lorenzetti L, Dinh N, van der Straten A, Fonner V, Ridgeway K, Rodolph M, et al. Systematic review of the values and preferences regarding the use of injectable pre-exposure prophylaxis to prevent HIV acquisition. J Int AIDS Soc. 2023;26 Suppl 2(Suppl 2):e26107.

13. Hennink M, Kaiser BN. Sample sizes for saturation in qualitative research: A systematic review of empirical tests. Social Science & Medicine. 2022;292:114523.

14. Hennink MM, Kaiser BN, Marconi VC. Code saturation versus meaning saturation: how many interviews are enough? Qualitative health research. 2017;27(4):591–608.

15. Lumivero. NVivo (Version 14) [Computer software]. 2023.

16. Nam-Sonenstein B, Sanders E. Why jails and prisons can’t recruit their way out of the understaffing crisis2024. Available from: https://www.prisonpolicy.org/blog/2024/12/09/understaffing/.

17. Bartholomew TS, Andraka-Cristou B, Totaram RK, Harris S, Doblecki-Lewis S, Ostrer L, et al. “We want everything in a one-stop shop”: acceptability and feasibility of PrEP and buprenorphine implementation with mobile syringe services for Black people who inject drugs. Harm Reduction Journal. 2022;19(1):133.

18. Shrestha R, Karki P, Altice FL, Dubov O, Fraenkel L, Huedo-Medina T, et al. Measuring Acceptability and Preferences for Implementation of Pre-Exposure Prophylaxis (PrEP) Using Conjoint Analysis: An Application to Primary HIV Prevention Among High Risk Drug Users. AIDS and Behavior. 2018;22(4):1228–38.

19. Vandergrift LA, Christopher PP. Do prisoners trust the healthcare system? Health Justice. 2021;9(1):15.

20. Walters SM, Frank D, Van Ham B, Jaiswal J, Muncan B, Earnshaw V, et al. PrEP Care Continuum Engagement Among Persons Who Inject Drugs: Rural and Urban Differences in Stigma and Social Infrastructure. AIDS and Behavior. 2022;26(4):1308–20.

21. Project THaR. Medicaid 1115 Reentry Waivers. 2025.

